# CLINICAL PROFILE AND OUTCOME OF SNAKE BITE ASSOCIATED ACUTE KIDNEY INJURY- A RETROSPECTIVE STUDY

**DOI:** 10.64898/2026.01.15.26343684

**Authors:** E Mahesh, Mohammed Yousuff, N Monika, Pooja Prabhu, M S Gireesh, R Rajashekar, V Hamsa

## Abstract

**Background:** Snakebite is a neglected tropical disease with a high burden in South Asia, particularly India. Acute kidney injury (AKI) is one of the most serious complications of snake envenomation, which has significant morbidity, mortality and risk of chronic kidney disease (CKD). The present study aimed to evaluate the incidence, predictors, and outcomes of snakebite-associated AKI (SBE-AKI) in a tertiary care centre.

**Methods:** We retrospectively analysed **325 patients** with snakebite envenomation, admitted to our institution. Demographic, clinical, laboratory, and treatment variables were compared between patients with and without AKI. AKI was staged according to KDIGO criteria. Renal biopsy was performed in selected patients. Outcomes assessed included recovery, Progression to CKD, and mortality.

**Results:** Of the 325 patients, **79 (32.1%) developed AKI**. Patients with AKI were significantly younger (mean age 34 vs. 45 years, *p* = 0.001). Delay in anti-snake venom (ASV) administration (18 vs. 6 hrs, *p* = 0.001), need for inotropes (41.8% vs. 14.2%, *p* = 0.001), and mechanical ventilation (36.7% vs. 6.9%, *p* = 0.001) were strong predictors. Proteinuria was more frequent in AKI (80% vs. 32.5%). Among AKI patients, 57% had stage 3 AKI; 39.2% required dialysis. Biopsy (n=8) showed acute tubular necrosis in 37.5% and cortical necrosis in 25%. Outcomes included **77.2% recovery, 6.3% progression to CKD, and 16.5% mortality**.

**Conclusion:** SBE-AKI is a common and serious complication of snakebite. Delay in ASV administration, hemodynamic instability, proteinuria, advanced AKI stage and cortical necrosis predict poor outcomes. Early ASV, timely dialysis, and long-term nephrology follow-up are essential to improve survival and reduce CKD progression.

## INTRODUCTION

Snakebite envenomation is a major yet neglected tropical disease, affecting rural and agricultural communities in Asia, Africa, and Latin America. The World Health Organization (WHO) estimates that 4.5–5.4 million people are bitten annually worldwide, resulting in 1.8– 2.7 million cases of envenomation and 81,000–138,000 deaths each year^1^. India contributes nearly half of this global burden, with an estimated 58,000 deaths annually, accounting for up to 0.5% of all deaths in the country^2^. Snakebite is thus not only a medical emergency but also a significant public health and occupational hazard in developing nations.

Acute kidney injury (AKI) is one of the most serious systemic complications of snakebite. The incidence of snakebite-associated AKI (SBE-AKI) varies widely, ranging from 13–32% in India and up to 45% in Southeast Asia, depending on the offending species, geographic distribution, and timeliness of treatment^3^□□. Mortality in patients with SBE-AKI remains high, between 20–50% in earlier studies□. Moreover, even among survivors, there is an appreciable risk of progression to chronic kidney disease (CKD), thereby contributing to the global CKD burden^12^.

The pathogenesis of SBE-AKI is multifactorial. Hemotoxic and myotoxic venoms from species such as *Viperidae* and *Elapidae* cause systemic and renal injury via multiple mechanisms, including: Direct nephrotoxicity of venom components (phospholipase A2, metalloproteinases, serine proteases). Hemodynamic instability from shock and capillary leak. Coagulopathy and thrombotic microangiopathy, leading to cortical necrosis.

Pigment nephropathy due to hemoglobinuria and myoglobinuria from intravascular hemolysis and rhabdomyolysis. Secondary sepsis and local tissue necrosis, further aggravate renal insult□□□.

Histologically, the renal lesions range from acute tubular necrosis (ATN), the most frequent and potentially reversible lesion, to acute interstitial nephritis (AIN) and diffuse cortical necrosis, which is associated with irreversible renal failure□□^11^□.

Access to anti-snake venom (ASV) and renal replacement therapy (RRT) are central to reducing mortality□. Unfortunately, in rural India and other resource limited regions, delays in ASV administration and limited availability of dialysis often lead to poor outcomes^3^□□.

Given this background, the present study was undertaken to evaluate the clinical profile, predictors, and outcomes of SBE-AKI in a tertiary care setting.

## OBJECTIVES

1. To evaluate the clinical profile of snake bite patients.
2. To determine predictors of developing AKI following snake bite.
3. To assess outcomes of patients with snakebite-related AKI.

## METHODOLOGY

This was a retrospective observational study conducted at hospitals attached to Ramaiah University of Applied Sciences, Bangalore. All patients with history of snakebite were included. AKI was diagnosed and staged according to KDIGO guidelines. Patients were grouped into AKI and non-AKI cohorts. Clinical, laboratory, and outcome parameters were compared between groups.

## STATISTICAL ANALYSIS

Data entry was done in Microsoft Excel. Statistical analysis was performed using SPSS version 21.

Descriptive statistics of the explanatory and outcome variables were calculated by mean, standard deviation for quantitative variables, frequency and proportions for qualitative variables.

Inferential statistics like Chi-square test was applied for qualitative variables to find the association. Independent sample t test was applied to compare the quantitative parameters between the groups. The level of significance was set at 5%

## RESULTS

A total of 325 patients with snakebite were included in the study, of whom 79 (32.1%) developed acute kidney injury (AKI).

### Demographic characteristics

**TABLE 1.** COMPARISON OF THE PAREMETERS BETWEEN THE GROUPS.

|  |  | Groups |  | p value |
| --- | --- | --- | --- | --- |
|  |  | Non-AKI | AKI |  |
| Age |  | 45.34 ± 16.64 | 34.17 ± 17.37 | 0.001* |
| Gender | Females | 61 (24.8%) | 19 (24.1%) | 0.893 |
|  | Males | 185 (75.2%) | 60 (75.9%) |  |
| DM | Absent | 231 (93.9%) | 69 (87.3%) | 0.057 |
|  | Present | 15 (6.1%) | 10 (12.7%) |  |
| HTN | Absent | 230 (93.5%) | 68(86.1%) | 0.038* |
|  | Present | 16 (6.5%) | 11 (13.9%) |  |

The mean age of patients in the AKI group was significantly lower than in the non-AKI group with male preponderance(34.2 ± 17.4 vs. 45.3 ± 16.6 years, *p* = 0.001). Hypertension was significantly more frequent among AKI patients (13.9% vs. 6.5%, *p* = 0.038), while diabetes mellitus showed a non-significant trend towards higher prevalence in AKI patients (12.7% vs. 6.1%, *p* = 0.057).

### Clinical parameters

Patients who developed AKI had a significantly higher requirement for inotropes (41.8% vs. 14.2%, *p* = 0.001) and mechanical ventilation (36.7% vs. 6.9%, *p* = 0.001) compared with the non-AKI group. Urine output at presentation differed markedly: oliguria was seen in 32.9% and anuria in 12.7% of AKI patients, whereas almost all non-AKI patients had normal urine output (*p* = 0.001).

The time interval from bite to ASV administration was significantly longer in the AKI group (18.1 ± 18.3 hrs) compared with non-AKI patients (6.1 ± 6.0 hrs, *p* = 0.001). The total ASV dose required was also higher in the AKI group (20.4 ± 7.7 vials vs. 12.8 ± 10.6 vials, *p* = 0.001). Mean hospital stay was nearly three times longer in AKI patients (11.1 ± 9.5 vs. 4.7 ± 2.6 days, *p* = 0.001).

**TABLE 2.**
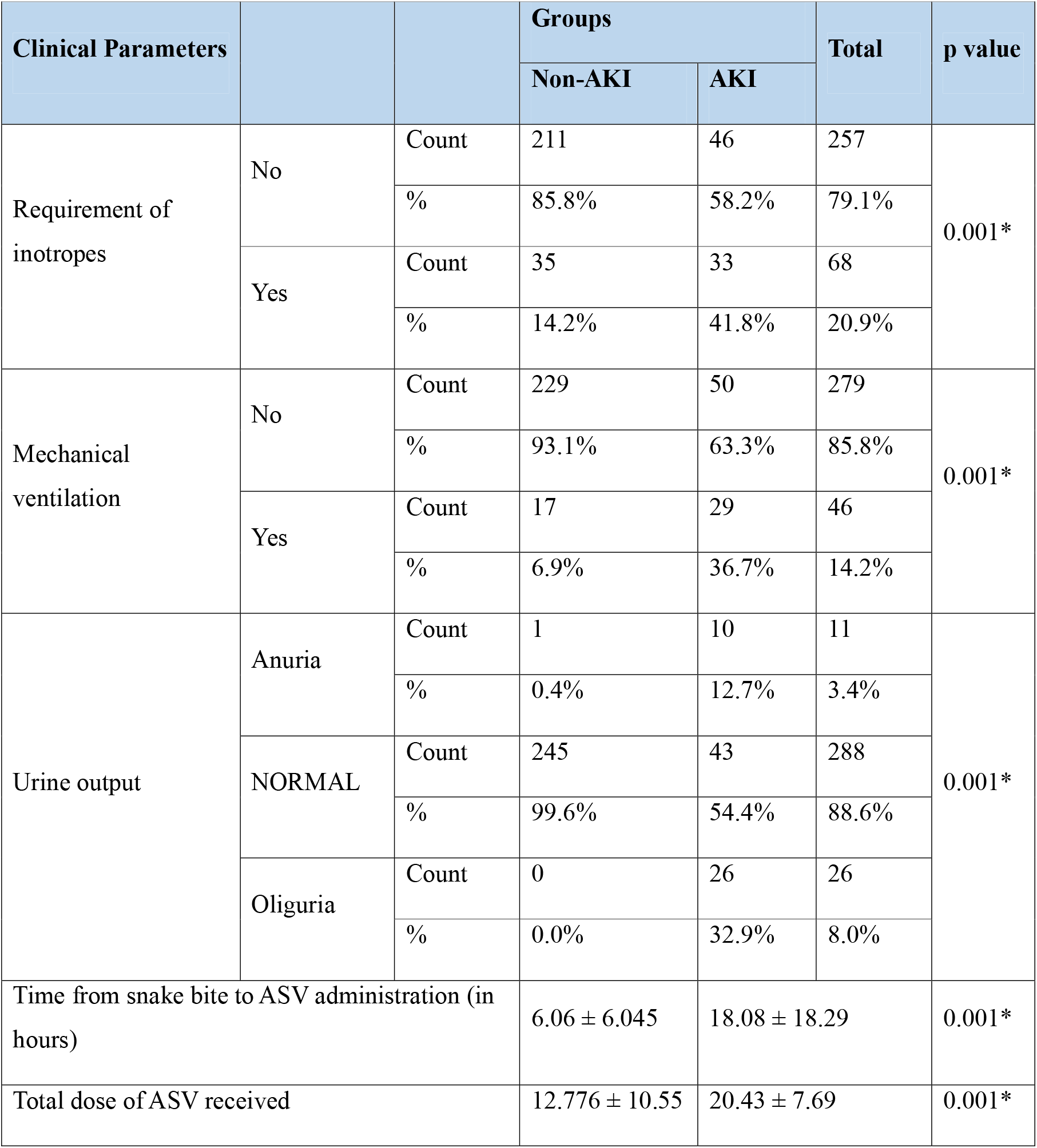
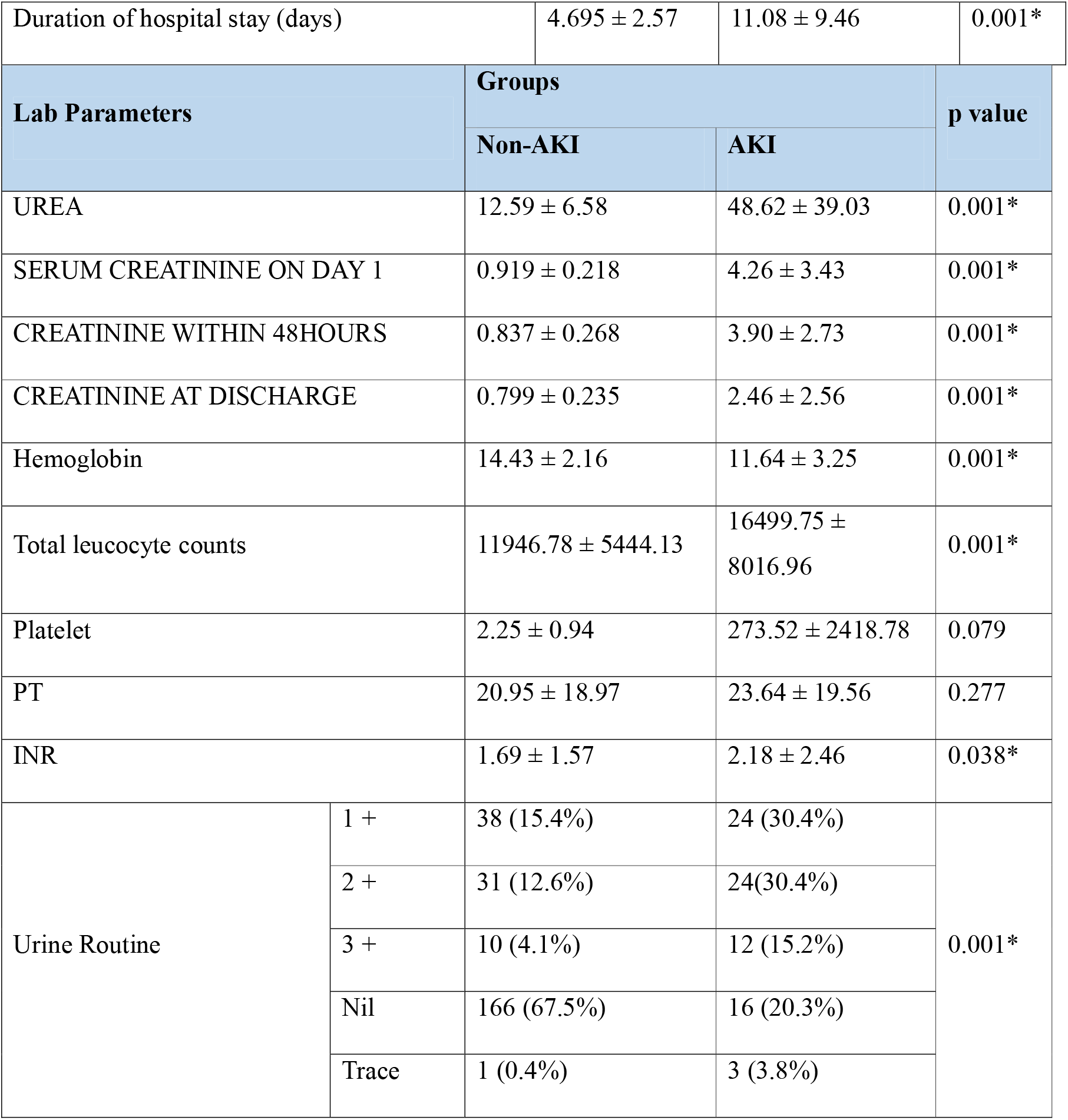
COMPARISON OF THE CLINICAL PARAMETERS AND LABORATORY BETWEEN THE GROUPS.

### Laboratory findings

Patients with AKI had markedly elevated serum urea (48.6 ± 39.0 vs. 12.6 ± 6.6 mg/dL, *p* = 0.001) and serum creatinine at day 1 (4.26 ± 3.43 vs. 0.92 ± 0.22 mg/dL, *p* = 0.001). Anemia was significantly more common in the AKI group (Hb 11.6 ± 3.3 vs. 14.4 ± 2.2 g/dL, *p* = 0.001), as was leukocytosis (TLC 16,499 ± 8,017 vs. 11,947 ± 5,444 cells/mm^3^, *p* = 0.001). INR values were also higher in the AKI group (2.18 ± 2.46 vs. 1.69 ± 1.57, *p* = 0.038). Urinalysis abnormalities (proteinuria) were significantly more frequent among AKI patients (80% vs. 32.5%, *p* = 0.001).

### Complications and interventions

Compartment syndrome occurred in 38% of AKI patients compared to 14.2% of non-AKI patients (*p* = 0.001). Surgical interventions such as fasciotomy (29.1% vs. 5.7%) and debridement (25.3% vs. 16.3%) were more common in the AKI group. Infective complications were also significantly higher in AKI patients, with cellulitis in 54.4% (vs. 22.8% in non-AKI), necrotizing fasciitis in 3.8% (vs. 0.4%), and cases of wet gangrene and DIC with MODS occurring exclusively in the AKI group.

**TABLE 3.** DISTRIBUTION OF THE PARAMETERS IN AKI GROUP AKI subgroup analysis.

AKI subgroup analysis
|  |  | Frequency | Percent |
| --- | --- | --- | --- |
| URINE OUTPUT | Anuria | 10 | 12.7 |
|  | Normal | 43 | 54.4 |
|  | Oliguria | 26 | 33 |
| STAGE OF AKI | 1.0 | 23 | 29.1 |
|  | 2.0 | 11 | 13.9 |
|  | 3.0 | 45 | 57.0 |
| RRT | 0 | 48 | 60.8 |
|  | Yes | 31 | 39.2 |
| BIOPSY | 0 | 71 | 89.9 |
|  | Acute on chronic interstitial nephritis | 2 | 25 |
|  | ATN | 3 | 37.5 |
|  | ATN with acute on chronic interstitial nephritis | 1 | 12.5 |
|  | Complete cortical necrosis | 2 | 25 |
| OUTCOME | AKI recovered | 61 | 77.2 |
|  | Death | 13 | 16.5 |
|  | Progressed to CKD | 5 | 6.3 |

Within the AKI cohort, stage 3 AKI was most common (57%), followed by stage 1 (29.1%) and stage 2 (13.9%). RRT was required in 39.2% of AKI patients, predominantly hemodialysis.

Renal biopsy was performed in 8 patients: ATN was the most frequent lesion (37.5%), followed by acute on chronic interstitial nephritis, and complete cortical necrosis (25%).

### Outcomes

Of the AKI patients, 77.2% recovered renal function completely, 6.3% progressed to CKD, and 16.5% succumbed to illness. Mortality was strongly associated with stage 3 AKI, anuria, shock, and delayed ASV administration.

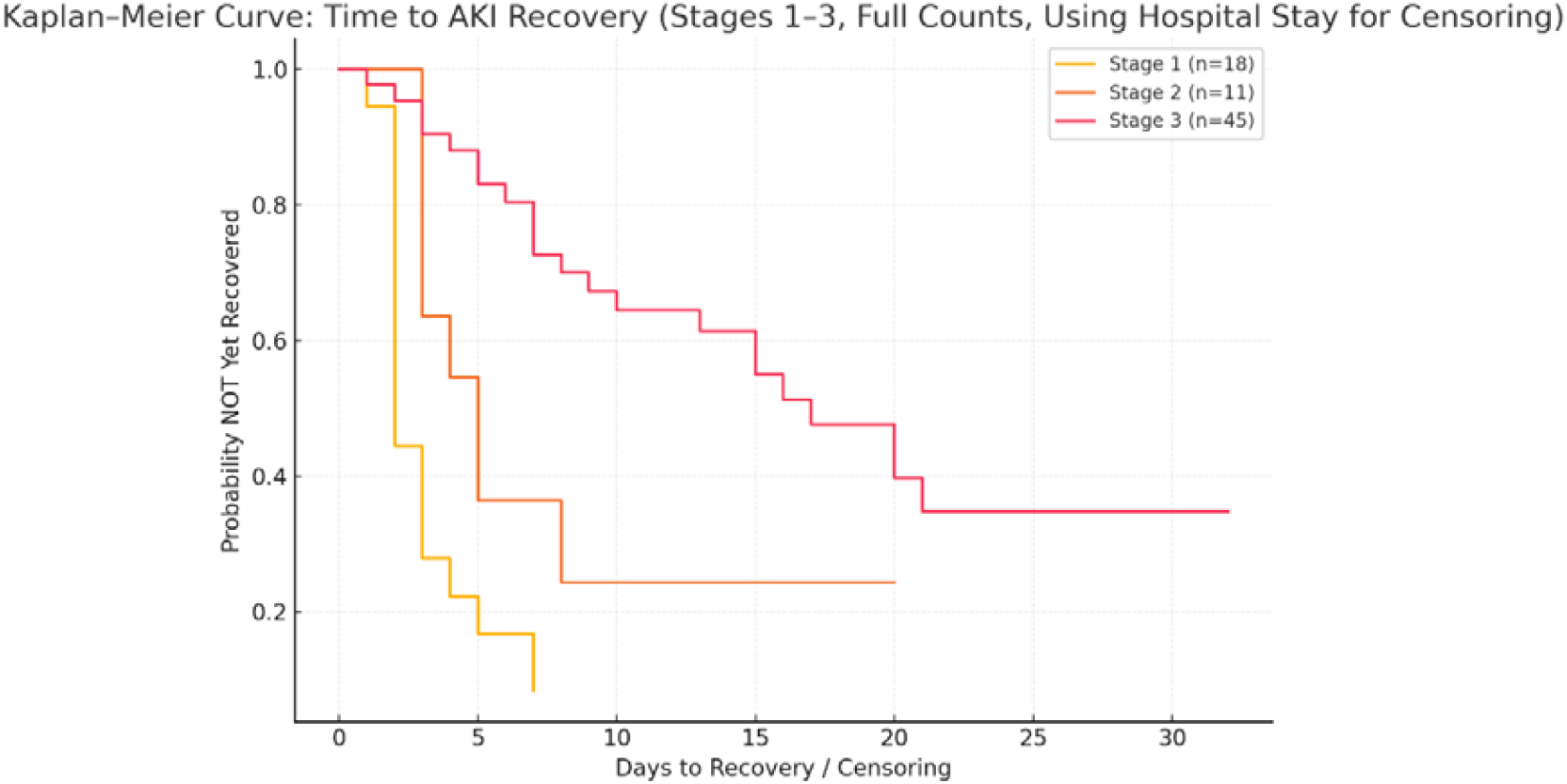

Kaplan meier curve shows the trend in recovery of AKI based on AKI severity. Rapid recovery was seen in stage 1 AKI where recovery was seen by 5-7 days. AKI stage 3 shows longest recovery time indicating more severe injury with delayed renal recovery.

**TABLE 4.** MULTIVARIATE REGRESSION.

| Predictor | AKI Severity (Ordinal) OR (95% CI) | p-value | Dialysis Requirement (Binary) OR (95% CI) | p-value | Death vs Recovery (Multinomial) OR (95% CI) | p-value |
| --- | --- | --- | --- | --- | --- | --- |
| Age (per year) | 1.00 (0.97–1.03) | 0.989 | 1.05 (1.01–1.10) | 0.026* | 0.98 (0.93–1.04) | 0.456 |
| ASV Delay (per hour) | **1.04 (1.01–1.08)** | **0.016\*** | **1.06 (1.02–1.09)** | **0.002\*** | 1.02 (0.97–1.06) | 0.490 |
| ASV Dose | 0.98 (0.91–1.05) | 0.516 | 0.99 (0.80–1.20) | 0.753 | 1.07 (0.96–1.19) | 0.244 |
| Male Sex | 1.02 (0.30–3.00) | 0.972 | 0.93 (0.25–3.40) | 0.920 | 0.30 (0.04–2.10) | 0.225 |
| Diabetes (Absent) | 5.00 (0.92–10.0) | 0.063 | 7.68 (0.92–12.0) | 0.062 | 3.66 (0.18–73.8) | 0.397 |
| Hypertension (Absent) | 0.25 (0.05–1.50) | 0.124 | 0.95 (0.05–1.50) | 0.959 | 0.14 (0.01–1.91) | 0.139 |
| Inotropes (No) | 0.97 (0.30–1.80) | 0.957 | 1.63 (0.50–3.50) | 0.497 | 0.27 (0.04–1.69) | 0.161 |
| Mechanical Ventilation (No) | **0.17 (0.05–0.61)** | **0.007\*** | **0.08 (0.02–0.33)** | **0.001\*** | **0.08 (0.01–0.58)** | **0.013\*** |

Predictors of AKI Severity (Ordinal Logistic Regression): Every hour delay increases odds of more severe AKI by 4.2%. Not requiring ventilation reduces odds of severe AKI by 83%.

Predictors of Dialysis Requirement (Binary Logistic Regression):

Predictors of Mortality (Multinomial Logistic Regression): Mechanical ventilation is the **single strongest predictor of death** in snakebite AKI.

## DISCUSSION

### Demographic and Clinical Predictors

The majority of patients in our study were young males, consistent with the occupational risk of farming and outdoor activities. Male predominance has been consistently reported across Indian studies□□□. The mean age was significantly lower in AKI patients (34 years) compared with non-AKI (45 years), reinforcing that snakebite nephropathy primarily affects healthy, economically active populations.

Pre-existing hypertension was significantly more common in the AKI group, and diabetes showed a trend towards higher prevalence. While not traditionally considered risk factors in envenomation, these comorbidities may reduce renal reserve, thereby amplifying the impact of venom-induced ischemia and coagulopathy.

Clinically, the need for inotropes and mechanical ventilation correlated strongly with AKI, underscoring that systemic severity of envenomation predicts renal outcomes. Reduced urine output (oliguria/anuria) was an early clinical sign of impending renal failure, consistent with previous reports□.

### The Role of ASV: Timing and Dose

A critical finding was the delay in ASV administration. AKI patients received ASV at a mean of 18 hours post-bite, compared with only 6 hours in non-AKI. This threefold delay was highly significant and highlights the time-sensitive nature of ASV therapy. Early administration within 4–6 hours is crucial for neutralizing circulating venom, preventing systemic complications, and reducing AKI risk□□□.

Additionally, AKI patients required a higher total ASV dose (20 vials vs. 13 vials), likely reflecting greater venom load and severity of envenomation.

### Laboratory Predictors and Urinalysis

Our AKI cohort demonstrated classical biochemical derangements: markedly elevated serum urea and creatinine, along with significant anemia, leukocytosis, and prolonged INR. These reflect hemolysis, systemic inflammation, and venom-induced coagulopathy.

A notable observation was the strong association between AKI and urinary abnormalities, present in 80% of AKI patients vs. 32.5% of non-AKI. Proteinuria suggests glomerular barrier injury, while hematuria may arise from both glomerular and tubular lesions. Prior studies□□^1^□ have correlated these findings with renal biopsy features such as acute tubular necrosis (ATN), interstitial nephritis, and cortical necrosis. Importantly, urinalysis is a simple, low-cost bedside tool that may help in early risk stratification, even before overt azotemia develops.

### Staging of AKI and Prognostic Value

Using KDIGO criteria, stage 3 AKI was the most common (57%), followed by stage 1 (29%) and stage 2 (14%). This indicates that most patients present late with advanced renal injury, often necessitating dialysis. In our cohort, nearly 40% required RRT, similar to other Indian studies□□□.

Staging also predicted outcomes: patients with stage 1 AKI had higher rates of recovery, while stage 3 was strongly associated with mortality and CKD progression.

### Histopathology of Snakebite Nephropathy

Renal biopsy, though performed in a small subset, provided valuable insights. ATN was the predominant lesion, confirming it as the morphological hallmark of snakebite AKI□□^1^□. ATN results from a combination of ischemia, pigment nephropathy (hemoglobin/myoglobin), and direct tubular cytotoxicity of venom enzymes such as phospholipase A2.

Cortical necrosis, observed in 25% of biopsied patients, is less common but carries grave implications, usually leading to irreversible renal failure. Acute interstitial nephritis (AIN) was also noted, reflecting immune-mediated injury or hypersensitivity to venom proteins. Importantly, AIN may respond to corticosteroid therapy, suggesting a potential therapeutic role for early renal biopsy in select patients.

### Outcomes and Progression to CKD

Encouragingly, 77% of AKI patients recovered renal function. However, 16.5% died and 6.3% progressed to CKD. Mortality was strongly associated with stage 3 AKI, anuria, shock, and delayed ASV administration. CKD progression occurred predominantly in patients with cortical necrosis or prolonged dialysis requirement.

This finding aligns with the emerging concept of the AKI-to-CKD continuum, where survivors of severe AKI remain at risk of long-term renal impairment^11^. Snakebite nephropathy, therefore, contributes not only to acute mortality but also to the burden of chronic kidney disease in endemic regions^12^.

Our findings align with prior Indian and Southeast Asian studies□□□,^11^□□^11^, which emphasize: 1) Early ASV as the cornerstone of prevention. 2) ATN as the commonest lesion, cortical necrosis as the worst prognosis. 3) High recovery rates if dialysis is available. 4) Significant risk of CKD in survivors.

Across all multivariable models, time from snakebite to ASV administration and requirement of mechanical ventilation emerged as the strongest and most consistent predictors of adverse renal outcomes. Delay in ASV increased the odds of severe AKI and dialysis requirement, while the need for mechanical ventilation reflecting systemic envenomation severity was the most powerful predictor of both AKI severity and mortality. Age mildly increased dialysis risk. No significant associations were found with sex, diabetes, hypertension, or total ASV dose.

## CONCLUSION

Snakebite is an important and preventable cause of AKI in tropical countries, disproportionately affecting young rural populations. Delay in ASV administration, presence of shock and respiratory failure, oliguria/anuria, and severe local complications were significant predictors of AKI.

While most patients recovered, cortical necrosis and severe AKI led to CKD progression or death in a significant proportion. Early ASV administration, dialysis availability, and long-term follow-up are essential to reduce morbidity and mortality from SBE-AKI.

## Data Availability

All data produced in the present study are available upon request to the authors

## LIMITATIONS

This was a Single-center study.

Renal biopsy performed in only a small proportion of patients.

No uniform protocol for ASV dosing.

Long-term CKD outcomes not available in all patients.

